# Household behaviour change interventions to improve sanitation and hygiene practices in urban settings: a scoping review

**DOI:** 10.1101/2024.08.20.24312313

**Authors:** Clara MacLeod, Katherine Davies, Mwamba M Mwenge, Jenala Chipungu, Oliver Cumming, Robert Dreibelbis

## Abstract

**Introduction:** Behaviour change interventions have the potential to improve sanitation and hygiene practices in urban settings. However, the evidence on which behaviour change interventions are effective is unclear. This scoping review assesses the effectiveness of behaviour change interventions on sanitation and hygiene practices in urban settings.

**Methods:** We performed electronic searches across five databases and one grey literature database to identify relevant studies published between 1 January 1990 and 20 November 2023 in English. Eligible study designs included randomised and non-randomised controlled trials with a concurrent control. Studies were eligible for inclusion if they reported a behaviour change intervention for improving sanitation and/or hygiene practices in an urban setting. Individual behaviour change intervention components were mapped to one of nine intervention functions of the capabilities, opportunities, motivations, and behaviour (COM-B) framework. Risk of bias was assessed for each study using an adapted Newcastle-Ottawa scale.

**Results:** After de-duplication, 8,249 documents were screened by abstract and title, with 79 documents retrieved for full-text screening. We included 13 studies ranging from low- to high-quality. The behaviour change interventions had mixed effects on sanitation and hygiene practices in urban settings. Specifically, interventions improved latrine quality but not safe child faeces disposal. Interventions often improved handwashing with soap at key times and sometimes increased the presence of soap and water at the handwashing facility. There is limited evidence on the effect on food hygiene practices. Most study outcomes were measured between 6 and 12 months after intervention implementation, which may undermine the sustainability of behaviour change interventions.

**Conclusion:** Despite mixed effects on sanitation and hygiene outcomes, behaviour change interventions can improve certain practices in urban settings, such as latrine quality improvements and handwashing with soap at the household or compound level. More ambitious behaviour change interventions are needed to reduce disparities in sanitation and hygiene access in urban areas globally.

## Introduction

Addressing sanitation and hygiene in urban areas, particularly in informal settlements, is essential for achieving Sustainable Development Goal (SDG) targets on water, sanitation, and hygiene (WASH) (SDG target 6.2). Lack of access to sanitation and hygiene is associated with enteric (1) and respiratory infections (2). In 2022, an estimated 36% of urban residents did not have access to safely managed sanitation, 25% of whom had access to basic sanitation, as defined by the WHO and UNICEF Joint Monitoring Programme (JMP) (3). It was also estimated that 17% of urban residents did not have access to basic hygiene services, with 10% having access to a limited hygiene facility and 7% with no access at all in 2022 (3). However, urban coverage of safely managed sanitation and basic hygiene services varied between countries and regions. Access to basic hygiene services was lowest in sub-Saharan Africa and Oceania and lowest in sub-Saharan Africa for access to safely managed sanitation (3). There are also marked sub-national disparities in access to sanitation and hygiene between high-income and low-income urban areas. Informal settlements, where approximately one-quarter of the global urban population resides, often lack formal WASH services (4). In the urban United States, in 2019, almost one million persons lacked access to at least basic sanitation, especially among people experiencing homelessness and substandard housing (5).

Safe sanitation and hygiene practices, such as latrine use, safe handling and disposal of faeces, handwashing with soap at key moments (e.g., after using the toilet, before food preparation), and hygienic food preparation and storage practices, are important for the prevention of communicable diseases, especially in urban areas where population densities are high. Two previous reviews found that behaviour change interventions achieved mixed results for improving WASH behaviours, such as handwashing with soap (6,7). However, these reviews did not disaggregate results between urban and rural settings. An understanding of behaviour change interventions that have been implemented specifically in urban settings can inform more effective future interventions, as well as identify areas for future research.

Behavioural frameworks and theories have been used to develop and design interventions to target sanitation and hygiene behaviours. Specifically, they can be used to understand the various factors that drive behaviour. For example, Michie et al. (8) developed a behavioural framework, known as the capabilities, opportunities, motivations, and behaviour (COM-B) framework (8), widely used in behaviour change programming. The COM-B framework identifies nine intervention functions that can be used to identify determinants of behaviour (8). In this scoping review, we identify and map the behaviour change intervention components used in the included studies to the nine intervention functions of the COM-B framework and assess their effectiveness, individually or in combination, on targeted sanitation and hygiene behaviours.

The aim of this scoping review is to evaluate the effectiveness of behaviour change interventions on household-level sanitation and hygiene practices in urban settings. The objectives are to: 1) identify household-level behaviour interventions targeting sanitation and hygiene practices in urban settings, 2) map the intervention components to the COM-B framework intervention functions, 3) assess their effectiveness, and 4) identify evidence gaps for future research.

## Methods

This scoping review follows the five steps for scoping reviews outlined by Arksey and O’Malley (9). The five steps are summarised below in relation to this review. A scoping review, as opposed to a systematic review, was selected to explore the breadth of available literature and to iteratively search and review documents and extract relevant data. The protocol for this scoping review was pre-registered on OSF registries (https://osf.io/qghl). We used the Preferred Reporting Items for Systematic Reviews and Meta-Analyses extension for scoping reviews (PRISMAScR) guidelines (10). A PRISMAScR checklist is included in the Supplementary Information (Table A1).

### Step 1: Specify the research question

This scoping review seeks to answer the following question: “*what is the effectiveness of behaviour change interventions targeting household-level sanitation and hygiene practices in urban settings?”* The research question is deliberately broad to allow for a comprehensive mapping of behaviour change interventions to sanitation and hygiene outcomes.

### Step 2: Identify the relevant literature

We searched five databases to identify peer-reviewed literature: 1) PubMed, 2) Medline, 3) Global Health, 4) Cochrane Library, and 5) Web of Science. The grey literature search was conducted in the World Bank e-library database. Search terms related to behaviour change, sanitation and hygiene, and urban settings were combined with Boolean operators to search the databases, with search strategies adapted for each database. An example search strategy is published in the Supplementary Materials (Table A2). Searches were conducted on 20 November 2023. The search was limited to studies published in English and from 1 January 1990 onwards. The publication cut-off date was selected based on the introduction of the Millennium Development Goals in 1990 (MDG7: “To ensure access to drinking water and sanitation for all”). The reference lists of included studies and similar systematic reviews (6,7,11) were also hand-searched to identify additional relevant references.

### Step 3: Study selection

Only studies with interventions that sought to change behaviour evaluated against a concurrent control group were included. The comparison is the use of a programme with no promotional approach or other promotional approaches. Eligible study designs were randomised controlled trials (RCTs), including cluster-RCTs, quasi-RCTs, and non-randomised controlled trials and pre-post studies. There was no restriction on target population. We included individual, household, and community-level interventions with the specific aim to improve sanitation and/or hygiene practices in urban settings.

An urban setting is broadly characterised by high population density, the concentration of administrative bodies, infrastructure, and services, and income generation activities (12). The specific criteria for what constitute an urban setting varies by country and is usually defined by national governments. Urban areas also include informal settlements (13), slums (13), as well as people experiencing homelessness in unsheltered urban locations (5). We relied on author-reported definitions of an urban setting.

In this scoping review, we used the COM-B framework intervention functions to identify and classify the behaviour change approaches used in the included studies. The COM-B framework includes three domains and nine intervention functions, as defined by Michie et al. (8). The first domain, capability, is defined as having the necessary physical ability, stamina, skills (physical capability), or knowledge (psychological capability) to engage in the activities involved in performing a behaviour. Second, opportunity relates to factors that lie outside the individual and that influences one’s ability to perform a behaviour, such as physical opportunity (i.e., resource availability) or social opportunity (i.e., social norms). Third, motivation refers to the “brain processes that energize and direct behaviour” and can be triggered by fear or disgust, for example. The nine intervention functions, or broad categories of things one can do to change behaviour, include: i) education, ii) persuasion, iii) incentivisation, iv) coercion, v) training, vi) restriction, vii) environmental restructuring, xiii) modelling, and ix) enablement (Table 1) (8).

**Table 1.** Intervention function description developed by Michie et al. (2011) (**8**).

| Intervention function | Definition | Relevant COM-B domain(s) |
| --- | --- | --- |
| Education | Increasing knowledge and understanding by informing, explaining, showing and correcting | Capability & motivation |
| Persuasion | Using communication to induce positive or negative feelings or stimulate action | Motivation |
| Incentivisation | Creating an expectation of reward | Motivation |
| Coercion | Creating an expectation of punishment or cost | Motivation |
| Training | Increasing psychological or physical skills, or habit strength by explanation, demonstration, practice, feedback and correction | Capability & motivation |
| Restriction | Constraining behaviour by setting rules | Opportunity |
| Environmental restructuring | Constraining or promoting behaviour by shaping the physical or social environment | Opportunity |
| Modelling | Showing examples of the behaviour for people to imitate | Capability & motivation |
| Enablement | Providing support to improve ability to change in a variety of ways not covered by other intervention functions e.g. through medication, surgery, encouragement, moral support | Capability |

The two primary outcomes for this review were household sanitation and hygiene practices. Relevant sanitation outcomes included latrine use, latrine construction or rehabilitation, building a septic tank, lining a pit, safe faeces handling and disposal (including child faeces), connecting to a piped sewer network, formal safe pit emptying, and latrine cleanliness. Hygiene outcomes included those related to handwashing with soap, handwashing with soap at key times (e.g., before eating, before food preparation, after visiting the toilet, after children’s faeces disposal or cleaning the baby’s bottom, or other key times defined in the studies), and handwashing facility construction. Food hygiene outcomes included boiling or reheating food before eating, using safe drinking water to prepare food, and hygienic storage of food (e.g., food covered with lid or refrigeration).

All documents retrieved from electronic searches were transferred to Endnote for de-duplication. To identify relevant documents, three reviewers (CM, KD, and MM) screened documents by title and abstract, excluding only clearly irrelevant documents, i.e., not related to sanitation and hygiene behaviour change interventions and urban settings. Full texts of all potentially eligible documents were then retrieved and independently double-assessed for inclusion by three reviewers (CM, KD, and MM). Any disagreement between reviewers about eligibility following title and abstract screening was resolved through discussion to build consensus. Disagreement was resolved through discussion with a fourth reviewer (RD) where consensus could not be reached.

### Step 4: Extract, map, and chart the data

Study characteristics and results from included studies were double-extracted independently by three reviewers (CM, KD, and MM) using a standardised data extraction template in MS Excel and then cross-checked for accuracy. As with inclusion, a fourth reviewer (RD) provided arbitration if agreement on data extraction could not be reached. The data extraction form included information on study characteristics, such as author, publication date, study design, study dates, study location and urban setting, target population, and sample size. We also extracted data on behaviour change approaches and sanitation and hygiene outcomes (Supplementary Materials Table A4). Only results for intervention arms targeting sanitation and hygiene behaviours were extracted.

### Step 5: Summarise, synthesise, and report results

First, intervention components for each intervention arm were mapped to one of the nine intervention functions of the COM-B framework (8). Second, we recorded the measure of effect, 95% confidence interval, and p-value for each outcome. Third, we summarised the results of the included studies to describe the effect of the behaviour change interventions on the sanitation and hygiene outcomes. Finally, we identified evidence gaps.

### Risk of bias (quality) assessment

We assessed risk of bias in individual studies using an adapted Newcastle-Ottawa scale, as used in previous systematic reviews (1,2,8). The scale considered seven areas of bias: selection, response, follow-up, misclassification, outcome assessment, outcome measurement, and analysis bias. Each study received a score of up to nine, a higher score indicating a smaller risk of bias. Risk of bias was assessed by one reviewer (CM) for each study with a subset of scores reviewed by a second reviewer (KD). Any discrepancies between the two reviewers were resolved by a third reviewer (RD).

## Results

### Search results

Electronic searches were conducted on 20 November 2023, identifying 9,771 records. After removing duplicates, 8,218 records were screened by abstract and title. Thirty-one additional documents were identified through reference screening. Seventy-nine documents were sought for retrieval for full text screening. Thirteen documents were included in the review (Figure 1). The 65 documents excluded during full text screening are listed with reasons for exclusion in the Supplementary Materials (Table A5). Most studies excluded during full text review were either conducted in rural settings or did not have a control group.

**Figure 1.**
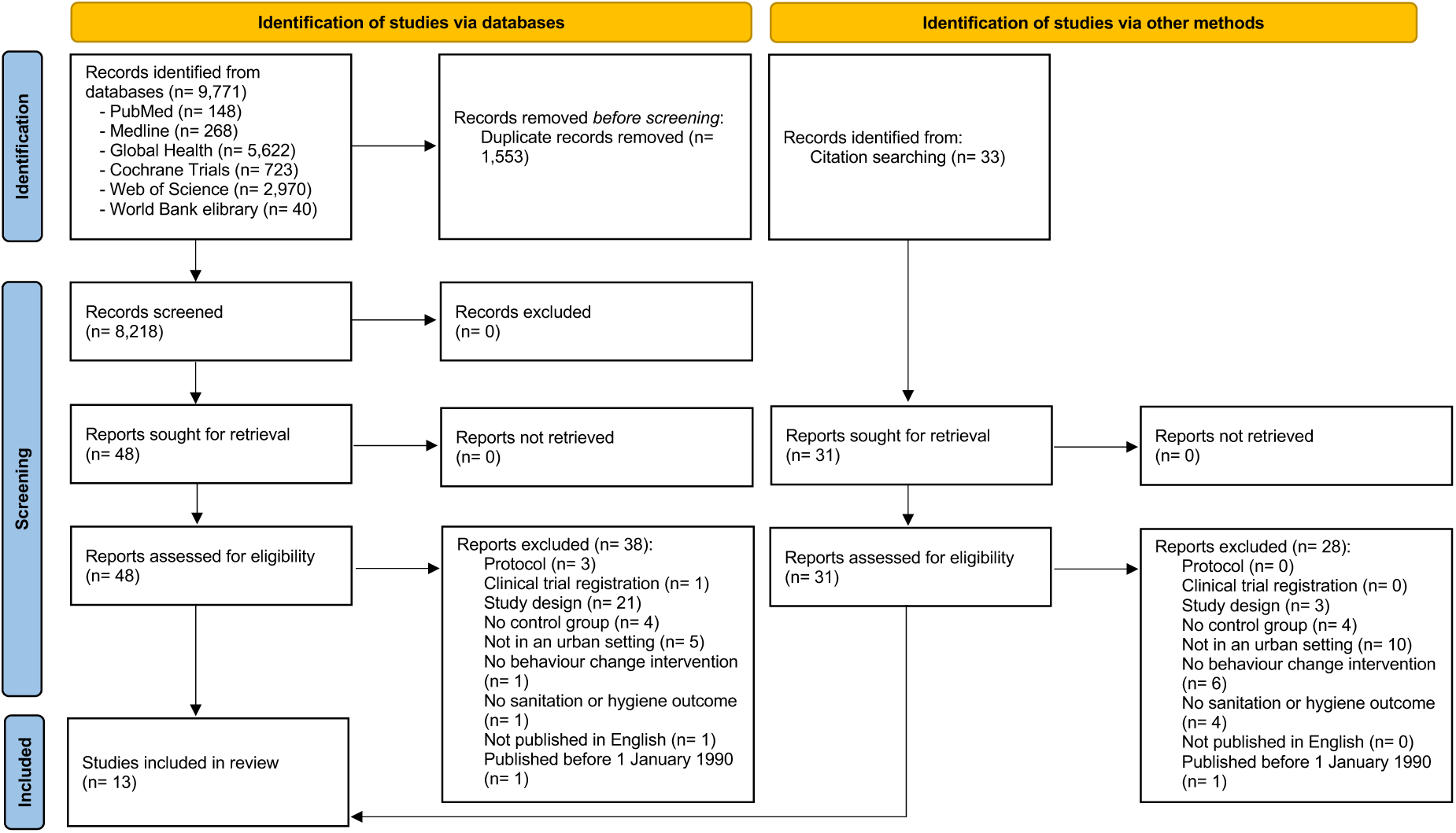
PRISMA flow chart.

### Description of included studies

The 13 included studies consisted of 12 peer-reviewed studies and 1 grey literature report (Table 1). Study designs included five cluster-randomised controlled trials (RCTs), three RCTs, three non-randomised trials, and two quantitative process evaluations. The two process evaluations were a controlled before-and-after study (14) and a cRCT (15). The studies were published between 2002 and 2022, though most studies were published after 2015. Almost all studies were conducted in sub-Saharan Africa (46%, n= 6) (1 in Côte d’Ivoire, 1 in Tanzania, 1 in Zambia, 1 in Uganda, 1 in Mozambique, and 1 in Kenya) or South Asia (46%, n= 6) (Bangladesh n= 3, Pakistan n= 2, and Nepal n= 1). One study was conducted in Latin America (Peru, n= 1). Eleven studies were in lower-middle-income countries (Bangladesh, Côte d’Ivoire, Tanzania, Kenya, Zambia, Pakistan, and Nepal), while two studies were in low-income countries (Mozambique and Uganda) and one in an upper-middle-income country (Peru). Seventy percent (n= 9) of studies were implemented in low-income urban areas, also called informal settlements, slums, or shanty towns among the included studies. One study was conducted in a commune within the city, while three were in peri-urban areas.

### Types of interventions

We identified 16 intervention arms across the 13 studies. The most frequently used COM-B framework intervention functions were environmental restructuring (n= 10), education (n= 10), and persuasion (n= 8) (Table 2). Most environmental restructuring was implemented in combination with other intervention functions (n= 10), such as education (n= 4), persuasion (n= 1), modelling (n= 1), and more than one intervention function (n= 4). Environmental restructuring alone was implemented in only one study (17). Two studies developed an intervention based on psychosocial theory, such as RANAS (24,25). Most studies compared interventions to a control group only (n= 10), while three studies compared two interventions to one another and to a control group. The interventions mainly targeted the individual level (54%, n= 7), such as users of shared sanitation or caregivers of children, while the other studies targeted the household level, for example, those with unimproved sanitation facilities (46%, n= 6). Thirty-one percent of studies (n= 4) included intervention components that were non- sanitation or hygiene related that were excluded from the scoping review analysis, e.g., received a cholera vaccine (18), water treatment (19,23), and water quality test results (20).

**Table 2.** Characteristics of included studies.

| Study | Year | Country | City | Urban setting | Study design | Follow up time point | Arms | Intervention functions (COM-B framework) by study arm | Intervention level | Number of participants (I/C) | Sanitation (outcome) | Hygiene (outcome) |
| --- | --- | --- | --- | --- | --- | --- | --- | --- | --- | --- | --- | --- |
| Alam et al. (16) | 2017 | Bangladesh | Dhaka | Informal settlement | cluster RCT | 6 months | 2 | A) Environmental restructuring + education<br>B) No intervention | Household | 1,214 households (609/605) | Latrine quality |  |
| Amon-Tanoh et al. (17) | 2021 | Côte d'Ivoire | Abidjan – Koumassi | City | cluster RCT | 5 months | 3 | A) Environmental restructuring + persuasion<br>B) Environmental restructuring only<br>C) No intervention | Compound | 73 compounds (23/25/25) |  | HW with soap |
| Bick et al. (14) | 2021 | Mozambique | Maputo | Informal settlement | Process evaluation | 12 months | 2 | A) Environmental restructuring + education<br>B) No intervention | Compound | 556 individuals (279/277) | Latrine quality | HW with soap |
| Biswas et al. (18) | 2012 | Bangladesh | Dhaka | Informal settlement | cluster RCT | 11 months | 2 | A) Environmental restructuring + persuasion + modelling<br>B) No intervention | Household | 400 households (100/200/100) |  | HW with soap |
| Bowen et al. (19) | 2012 | Pakistan | Karachi | Informal settlement | cluster RCT | 5 years | 2 | A) Environmental restructuring + persuasion + education<br>B) No intervention | Household | 461 households (160/141) |  | HW with soap |
| Davis et al. (20) | 2011 | Tanzania | Dar es Salaam | Peri-urban area | Non-randomised trial | Not stated | 2 | A) Education + persuasion<br>B) Education (comparison) | Household | 248 households (79/84/90/81) |  | HW with soap |
| Guiteras et al. (21) | 2015 | Bangladesh | Dhaka | Informal settlement | RCT | 3.5 months & 7 months | 3 | A) Education + persuasion + coercion<br>B) Environmental restructuring + modelling<br>C) Education (comparison) | Compound | 420 compounds (210/214) |  | HW with soap |
| Langford et al. (22) | 2013 | Nepal | Kathmandu | Informal settlement | Randomised trial | 6 months | 2 | A) Environmental restructuring + education + enablement<br>B) No intervention | Household | 88 households (45/43) |  | HW with soap |
| Luby et al. (23) | 2009 | Pakistan | Karachi | Informal settlement | cluster RCT | 18 months | 2 | A) Environmental restructuring + persuasion + education<br>B) No intervention | Household | 390 households (195/195) |  | HW with soap |
| Simiyu et al. (15) | 2022 | Kenya | Kisumu | Peri-urban area | Process evaluation | 2 months & 3.5 months | 2 | A) Environmental restructuring + persuasion + education<br>B) No intervention | Individual | 723 individuals (387/336) |  | HW with soap + food hygiene |
| Tidwell et al. (24) | 2019 | Zambia | Lusaka | Peri-urban area | RCT | 6 months | 2 | A) Persuasion + education + modelling + incentivisation + enablement<br>B) No intervention<br>+ elements of psychosocial theory | Individual | 928 individuals (474/454) | Latrine quality |  |
| Tumwebaze et al. (25) | 2015 | Uganda | Kampala | Informal settlement | Non-randomised trial | Not stated | 3 | A) Incentivisation<br>B) Incentivisation + enablement<br>C) No intervention | Household | 119 households (38/41/40) | Latrine quality |  |
|  |  |  |  |  |  |  |  | + elements of psychosocial theory |  |  |  |  |
| Yeager et al. (26) | 2002 | Peru | Lima | Informal settlement | Non-randomised trial | 12 months | 2 | A) Education<br>B) No intervention | Individual | 578 individuals (285/293) | Safe child faeces disposal |  |

### Outcomes

We identified 36 sanitation and hygiene outcomes across the 13 included studies. Fourteen outcomes related to sanitation and 21 to hygiene. Of the 21 outcomes related to hygiene, 19 were on hand hygiene and two were on food hygiene. The sanitation outcomes specifically related to latrine quality (n= 13) (14,16,24,25) and safe child faeces disposal (n= 1) (26). Hand hygiene outcomes included handwashing with soap and at various key moments (e.g., after using the toilet or before eating) (n= 13) (15,17,19,21–23) or the presence of soap and water at the household handwashing facility (n= 6) (14,18–21). The two food hygiene outcomes included using clean utensils for infant feeding and hygienic storage of leftover food (15).

Outcome definitions and measurement timepoints varied across the studies. Latrine quality was measured as either latrine cleanliness, such as no visible faeces in pan (16), having a rotation cleaning system (14,24), and cleaning frequency (25), or latrine privacy, measured as having an indoor and outdoor lock (14,24). Sanitation-related outcomes relied on both fieldworker observations (n= 8) and self-reporting (n= 5). Hygiene outcomes included either observed handwashing with soap (n= 12), self-reported handwashing with soap (n= 1), observed presence of a handwashing facility with soap and water (n= 6), and observed food hygiene practices (n= 2). However, each study measured handwashing with soap differently and often before or after different key moments. For example, one study reported daily handwashing rates (20), while another study reported the proportion of occasions participants washed their hands with soap after using the toilet (17). The observed presence of a handwashing with soap, often used as a proxy measure for handwashing, was measured consistently across studies, though measured at different timepoints after intervention implementation. Over half of the outcomes were measured between 6 months and 12 months after baseline (54%, n= 7), with two studies that measured outcomes less than six months after baseline (15,17). Two studies measured the medium and long-term effects of a behaviour change intervention on hygiene 18 months and 5 years post- intervention (19,23). Two studies did not state when they measured outcomes (20,25).

### Intervention effectiveness

There are mixed results on the effect of behaviour change interventions on sanitation and hygiene outcomes in urban settings. Interventions were associated with improved sanitation and hygiene practices in 27 out of the 36 outcomes, while there was no effect on 9 out of 36 outcomes. Interventions had a positive effect on 13 out of 14 outcomes related to sanitation. However, intervention effects on handwashing were mixed as 13 out of 20 outcomes had a positive effect and 7 out of 20 outcomes had no effect. The only two food hygiene outcomes included in this review had mixed results.

#### Effectiveness of sanitation interventions

Behaviour change interventions improved latrine quality (14,16,24,25), but did not improve safe child faeces disposal (26) (Table 3). Alam et al. (16) found that compounds that received a latrine cleaning intervention were significantly more likely to have cleaner toilets (e.g., no visible faeces on latrine pan) after the intervention compared to controls. Tidwell et al. (24) found intervention households were more likely to have cleaning rotas, inside and outside latrine locks, and toilets with simple covers or water seals. Tumwebaze et al. (25) found that shared toilet users that received either one of two latrine cleaning interventions were significantly more likely to clean their shared latrine. Bick et al. (14) found that compounds receiving an improved sanitation intervention were much more likely to be private, almost twice as likely to be observably clean, and twice as likely to be well-maintained. In addition, individual cleaning frequency was significantly higher and frequent collective cleaning was reported more often by intervention respondents. However, intervention compounds were unlikely to have and adhere to a formal rota for cleaning shared latrines. Yeager et al. (26) found that intervention respondents were not more likely to practice safe child faeces disposal than control respondents.

**Table 3.** Description of study results.

| Author | Intervention function by arm | Comparison | Outcome | Description of outcome | Effect measure | Results (95% CI and p-value where relevant) | Interpretation of results |
| --- | --- | --- | --- | --- | --- | --- | --- |
| **Sanitation** |  |  |  |  |  |  |  |
| Alam et al. (16) | A) Environmental restructuring + education | No intervention | Latrine quality | Visible faeces inside the pan (observed) | DID | -13% (95% CI -19%, -5%) | Compounds that received the intervention were significantly more likely to have cleaner toilets after the intervention. |
| Tidwell et al. (24) | A) Persuasion + education + modelling | No intervention | Latrine quality | Having a rotation cleaning system in place (reported) | RR | 1.16 (95% CI 1.05–1.30) (p=0.0011) | Plots that received the intervention were significantly more likely to have higher quality toilets across all four dimensions of quality improvement. |
|  |  |  |  | Having inside lock (observed) | RR | 1.34 (95% CI 1.10–1.64) (p=0.00081) |  |
|  |  |  |  | Having outside lock (observed) | RR | 1.27 (95% CI 1.06–1.52) (p=0.0028) |  |
|  |  |  |  | Toilets with simple covers or water seals (observed) | RR | 1.25 (95% CI 1.04–1.50) (p=0.0063) |  |
| Tumwebaze et al. (25) | A) Incentivisation | No intervention | Latrine quality | Shared sanitation users' cleaning behaviour (reported) | F-value (time) | 13.84 (p<0.005) | Shared toilet users that received either intervention were significantly more likely to have improved cleaning behaviour. |
|  | B) Incentivisation + restriction |  |  | Shared sanitation users' cleaning behaviour (reported) | F-value (time) | 14.71 (p<0.005) |  |
| Bick et al. (14) | A) Environmental restructuring + education | No intervention | Latrine quality | Have and adhere to a latrine cleaning rota (reported) | $\chi^2$ | 6.1 (p=0.013) | Intervention compounds were unlikely to have and adhere to a formal rota for cleaning shared latrines. However, individual cleaning frequency was significantly higher among intervention respondents compared to control respondents and frequent collective cleaning was reported more often by intervention respondents. In addition, intervention latrines were much more likely to be private, almost twice as likely to be observably clean, and twice as likely to be well-maintained. |
| | | | | Individual cleaning frequency (twice/week) | $\chi^2$ | 14 (p<0.001) | |
| | | | | Frequent collective cleaning (latrine cleaned on daily basis) | $\chi^2$ | 19 (p<0.001) | |
| | | | | Private latrine (working door and inside lock) (observed) | $\chi^2$ | 500 (p<0.001) | |
| | | | | Observably clean latrine (observed) | $\chi^2$ | 150 (p<0.001) | |
| | | | | Well-maintained latrine (slab/floor in good condition) (observed) | $\chi^2$ | 240 (p<0.001) | |
| Yeager et al. (26) | A) Education | No intervention | Safe faeces disposal | Safe child faeces disposal (observed) | Est diff | 0.002 | Intervention respondents were not more likely to practice safe child faeces disposal than control respondents. |
| **Hygiene** |  |  |  |  |  |  |  |
| Biswas et al. (18) | A) Environmental restructuring + persuasion | No intervention | Presence of soap and water | Presence of water and soap or soapy water at HWF (observed) | Proportion test | 60% [102/171] vs. 31% [28/90] (p<0.001) | Intervention respondents were significantly more likely to have water and soap or soapy water present at the hand washing place than control respondents. |
| Davis et al. (20) | A) Education + persuasion | B) Education (comparison) | Handwashing with soap | Handwashing rates (times per day) (reported) | t | 0.48 (0.34) | No significant differences between the intervention and control groups were observed in reported handwashing behaviours. |
| Bick et al. (14) | A) Environmental restructuring + education | No intervention | Presence of soap and water | Presence of a HWF with soap and water (observed) | X<sup>2</sup> | 0.12 (p= 0.729) | Few intervention latrines had signs of soap use at an HWF with no difference from control latrines. |
| Amon-Tanoh et al. (17) | A) Environmental restructuring + persuasion + enablement | No intervention | Handwashing with soap | Proportion of occasions during which hands washed with soap after using the toilet (observed) | aRR | 2.68 (95% CI 1.65–4.34) | The environmental restructuring, persuasion, and enablement intervention was effective at increasing handwashing with soap after toilet use, while environmental restructuring alone (provision of handwashing stations) had little effect. |
|  | B) Environmental restructuring |  |  |  | aRR | 1.89 (95% CI 1.16–3.08) |  |
| Guiteras et al. (21) | A) Education + persuasion + coercion | Education | Handwashing with soap | Handwashing after last defecation (used soap and water, both hands) (observed) | Est difference | 0.009 | The environmental restructuring and modelling intervention increased handwashing after last defecation and the presence of soap and water at the latrine.<br>The education, persuasion, coercion intervention did not increase handwashing after last defecation nor the presence of soap and water at the latrine. |
|  |  |  | Presence of soap and water | Presence of soap and water at latrine (observed) | DID | -0.068 |  |
|  | B) Environmental restructuring + modelling | No intervention | Handwashing with soap | Handwashing after last defecation (used soap and water, both hands) (observed) | Est difference | 0.048 (p<0.01) |  |
|  |  |  | Presence of soap and water | Presence of soap and water at latrine (observed) | DID | 0.540 (p<0.01) |  |
| Simiyu et al. (15) | A) Environmental restructuring + persuasion + education | No intervention | Handwashing with soap | HWS before infant food preparation (observed) | OR | 1.38 (95% CI 1.02–1.87) (p= 0.035) | The intervention improved handwashing with soap before food preparation and hygienic feeding of infants (i.e., using a utensil). However, the intervention had no effect on caregivers' handwashing practices before infant feeding and caregiver hygienic food storage practices. |
|  |  |  |  | HWS before infant feeding (observed) | OR | 0.92 (95% CI 0.68–1.25) (p= 0.6) |  |
|  |  |  | Food hygiene | Using a feeding utensil (observed) | OR | 3.5 (95% CI 1.91–6.56) (p= 0.00) |  |
|  |  |  |  | Hygienic food storage (observed) | OR | 0.00 (95% CI 0.00 -0.004) (p= 0.0001) |  |
| Luby et al. (23) | A) Environmental restructuring + persuasion + education | No intervention | Presence of soap and water | Presence of soap and water at latrine (observed) | RR | 79% vs. 53% (p= 0.001) | Mothers in households originally assigned to the intervention were 1.5 times more likely to have a place with soap and water to wash hands 18 months after the intervention. |
| Bowen et al. (19) | A) Environmental restructuring + persuasion + education<br>B) Persuasion | No intervention | Handwashing with soap | HWS before cooking (observed) | RR | 1.2 (95% CI 1.0–1.4) | Intervention households more commonly reported handwashing before cooking and before meals than control households five years after the intervention. |
|  |  |  |  | HWS before eating or feeding others (observed) | RR | 1.7 (95% CI 1.3–2.1) |  |
|  |  |  | Presence of soap and water | Presence of soap and water at latrine (observed) | Chi-square test | 97% [293/301] vs. 28% [45/159] (p<0.0001) | Intervention households were 3.4 times more likely to have a handwashing station with soap and water than control households five years after the intervention. * |
| Langford et al. (22) | A) Environmental restructuring + education | No intervention | Handwashing with soap | HWS after visiting the toilet (observed) | Chi-square test | 100% [45/45] vs. 91% [39/43] (p= 0.053) | Mothers in the intervention group were significantly more likely to wash hands with soap in four out of five key junctures: after cleaning a baby's bottom, before cooking, feeding the baby, and eating compared to the control group. |
|  |  |  |  | HWS after cleaning baby's bottom (observed) | Chi-square test | 100% [45/45] vs. 84% [36/43] (p= 0.005) |  |
|  |  |  |  | HWS before cooking (observed) | Chi-square test | 71% [32/45] vs. 2% [1/43] (p<0.001) |  |
|  |  |  |  | HWS before feeding the baby (observed) | Chi-square test | 62% [28/45] vs. 19% [8/43] (p<0.001) |  |
|  |  |  |  | HWS before eating (observed) | Chi-square test | 60% [27/45] vs. 0% [0/43] (p<0.001) |  |

HWS= handwashing with soap; RR= relative risk; aRR= adjusted relative risk; OR= odds ratio; DID= difference in difference
\* Results for both intervention arms are reported as one result versus the control

**Table 4.**
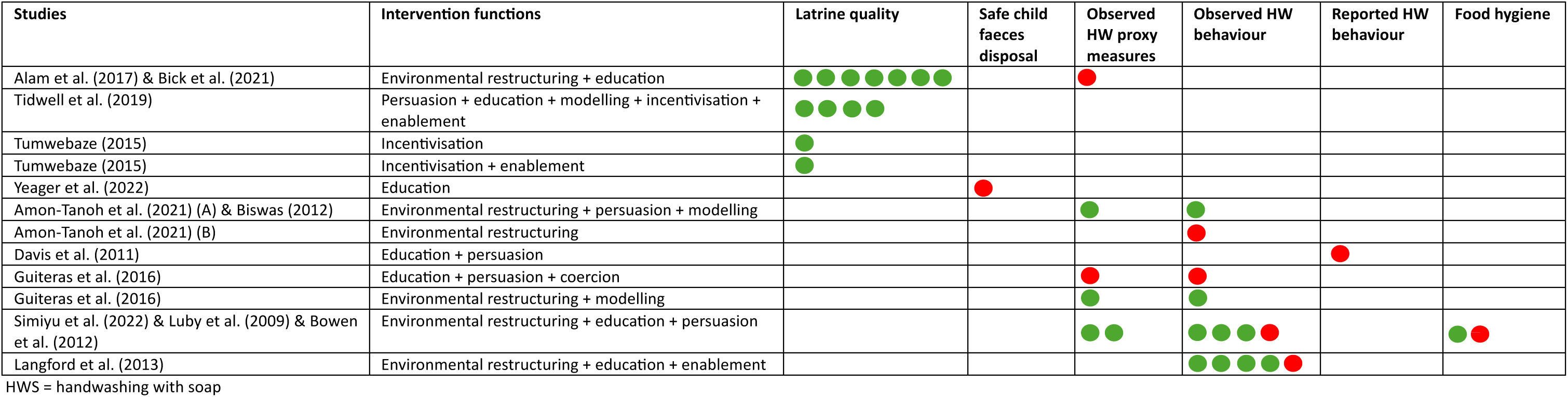
The effect of intervention functions, alone or in combination, on specific sanitation and hygiene outcomes. Each circle represents one outcome reported in an individual study. A red circle indicates a negative or no effect on the reported outcome, while a green circle indicates a positive effect on the reported outcome.

#### Effectiveness of hygiene

Behaviour change interventions had mixed results on improving hygiene practices in urban settings (Table 2).

### Observed presence of soap and water at the latrine

Behaviour change interventions sometimes increased the observed presence of soap and water at the handwashing facility. In Dhaka, Bangladesh, Biswas et al. (18) found that intervention respondents were significantly more likely to have water and soap or soapy water present at the hand washing place than control respondents. In another study in Dhaka, Bangladesh, one intervention arm (environmental restructuring and modelling) increased the presence of soap and water at the latrine, while another intervention arm (education, persuasion, coercion) did not (21). In Karachi, Pakistan, Luby et al. (23) followed up participants 18 months after a behaviour change intervention ended (27) and found that mothers in households originally assigned to the intervention were 1.5 times more likely to have a handwashing facility with soap and water. Five years after the same intervention followed up by Luby et al. (23), Bowen et al. (19) reported that intervention households were 3.4 times more likely to have a handwashing station with soap and water than control households. Bick et al. (14), found that few intervention latrines had signs of soap use at the household handwashing facility with no difference from control latrines in Maputo, Mozambique.

### Observed hygiene practices

Most behaviour change interventions improved observed handwashing with soap at key moments. Amon-Tanoh et al. (17) found that the ‘environmental restructuring, persuasion, and enablement’ intervention was effective at increasing handwashing with soap after toilet use, while environmental restructuring alone (provision of handwashing stations) had little effect. In Langford et al. (22), mothers in the intervention group were significantly more likely to wash hands with soap in four out of five key junctures compared to the control group: after cleaning a baby’s bottom, before cooking, feeding the baby, and eating. In Simiyu et al. (15), the intervention improved caregiver handwashing with soap before food preparation but had no effect on caregivers’ handwashing practices before infant feeding. In Bowen et al. (19), intervention households more commonly reported handwashing before cooking and before meals than control households five years after the intervention. In another study, one intervention arm (environmental restructuring and modelling) increased handwashing after last defecation, while another intervention arm (education, persuasion, and coercion) did not (21). The only study that investigated food hygiene practices found that the intervention improved observed hygienic feeding of infants (i.e., using a utensil) but had no effect on caregiver hygienic food storage practices (15) (Table 1).

### Reported hygiene practices

Only one study reported on handwashing behaviour. The study found no significant differences between the intervention and control groups were observed in reported handwashing behaviours (20).

### Risk of bias

The risk of bias ranged from high to low for sanitation and hygiene outcomes. In most studies, participants and those collecting the data could not be blinded to intervention allocation, but randomised study design, low loss to follow up, and blinding of data analysts contributed to higher Newcastle-Ottowa Scale scores. The self-reported or observation of outcomes, as well as high rates of loss to follow up, led to lower scores. Three studies were not randomised and seven had a loss to follow up >10%. Most sanitation and hygiene outcomes were observed. The full assessment is in the Supplementary Materials (Table A5).

## Discussion

This scoping review included 13 studies evaluating the effect of behaviour change interventions on sanitation and hygiene practices in urban settings. The results suggest that behaviour change interventions can improve certain sanitation and hygiene practices, such latrine quality and handwashing with soap at key moments. There is mixed and limited evidence on the effectiveness of behaviour change interventions on other outcomes, such as safe child faeces disposal and food hygiene practices. The 13 studies were implemented in 10 countries and primarily in urban informal settlements in sub-Saharan Africa and South Asia. No studies were conducted in high-income countries where disparities in access to sanitation and hygiene remain in urban areas. We also note that two excluded studies were implemented in both urban and rural settings but did not disaggregate results by setting (28,29). Compared to sanitation and hygiene interventions targeting rural areas, the evidence base is much more limited for the urban setting. Evidence specific to behaviour change interventions in urban settings is important for addressing the sanitation and hygiene challenges in this context.

Most studies included at least two intervention functions in their behaviour change interventions, thereby limiting the ability to tease out the specific effect from each intervention component. Almost all studies related to hand hygiene relied on environmental restructuring in combination with more traditional forms of interpersonal communication (e.g., persuasion, education, modelling). Among the environmental modification interventions, the majority focused on hardware provision, specifically providing an improved handwashing station to households. Nudges or environmental cues, which have generally shown to improve behavioural outcomes (30–32), warrant further exploration in urban settings. Other behaviour change approaches, such as community mobilisation, social marketing, advocacy, and financial incentives also warrant further exploration. For example, several studies have reported that willingness-to-pay for sanitation products and services is well under market prices in low-income urban areas (33–35). Financial incentives, such as income- or area-based subsidies, may bridge the gap between cost and willingness to pay for improved sanitation and safe emptying services (35).

The interventions targeted a narrow range of sanitation and hygiene behaviours. Latrine quality improvements were the most targeted sanitation-related behaviour. While sanitation quality can be an important predictor of sanitation use (36), latrine quality improvements alone may have limited impact for reaching SDG targets. Only one study evaluated safe child faeces disposal, and no studies targeted the use of latrines, safe pit emptying, or faecal sludge management. In addition, no studies evaluated the use of novel sanitation technologies designed for the urban marketplace, such as container-based sanitation (CBS). Handwashing with soap was the most targeted hygiene behaviour. Only one behaviour change intervention targeted food hygiene behaviours, which highlights an important evidence gap.

Outcome definitions and measurement timepoints varied significantly across the studies, thus making it difficult to compare results. For example, handwashing with soap was either measured via structured observation, self-reported behaviour, or proxy measures. Most included studies used structured observations of handwashing behaviour, often considered the gold standard for measuring behaviour., though more resource-intensive (37). Alternatively, some studies used the presence of a handwashing facility with soap and water as a proxy measure for handwashing behaviour. While this method allows for rapid and low-cost data collection, it’s accuracy may be limited (38). In addition, one study relied on self-reporting handwashing, which is prone to recall bias (37). Outcome measurement time points ranged from 6 to 12 months post-intervention, with only one study investigating long-term intervention effect. The findings suggest that behaviour change interventions were overall effective at improving certain sanitation and hygiene practices, but it is unclear whether they are effective long- term.

We note the limited scope of robust, large-scale interventions addressing sanitation and hygiene at the municipal or community-level. All interventions included in this review focused on household- or compound-level improvements. We considered compound-level interventions as household interventions as the interventions were delivered to a relatively small sample of households within clusters of compounds. No studies explicitly addressed community-level behaviour or behaviour change nor did they focus on connecting to municipal water or sewerage systems. With recent emphasis of urban sanitation programmes on Citywide Inclusive Sanitation (39), rigorous evaluation of efforts to improve urban sanitation are needed. While consistent water supply is necessary for safe sanitation and effective hygiene behaviours, only one included study adjusted for water supply in their analysis (24) and only one study provided information on water supply at baseline (16).

This scoping review has several limitations. First, our search was limited to English and may have missed relevant documents published in other languages. Second, we searched one grey literature database and may have missed additional relevant grey literature published elsewhere. Third, the outcomes amongst the included were too heterogeneous to conduct a meta-analysis. We also did not evaluate publication bias. Fourth, due to the nature of the interventions, blinding of participants and enumerators was often not possible, which may lead to outcome measurement bias. Results included in the review may also have been biased due to self-reporting of sanitation and hygiene outcomes. Finally, only one study evaluated food hygiene, which limits the generalisability of results.

## Conclusion

Our results suggest that behaviour change interventions have the potential to improve sanitation and hygiene practices in urban settings, such as latrine cleanliness and handwashing with soap at the household or compound level. However, more ambitious interventions should be evaluated to increase their impact. Opportunities for future interventions include evaluating community-level behaviour change interventions, connecting households to water or sewerage networks where available, CBS acceptability, uptake and use, and food hygiene practices. Nonetheless, this review highlights that behaviour change is an important component of interventions for sanitation and hygiene in urban settings.

## Conflict of interest

The authors have declared that no competing interests exist.

## Funding statement

This research was funded by the United Kingdom’s Foreign, Commonwealth and Development Office (grant code: 301186). The funder had no role in study design, data collection and analysis, decision to publish, nor preparation of the manuscript.

## Data Availability

All data produced in the present work are contained in the manuscript

## Supplementary materials

**Table A1.** The completed Preferred Reporting Items for Systematic reviews and Meta-Analyses extension for Scoping Reviews (PRISMA-ScR) checklist.

| SECTION | ITEM | PRISMA-ScR CHECKLIST ITEM | REPORTED ON PAGE # |
| --- | --- | --- | --- |
| **TITLE** |  |  |  |
| Title | 1 | Identify the report as a scoping review. | 1 |
| **ABSTRACT** |  |  |  |
| Structured summary | 2 | Provide a structured summary that includes (as applicable): background, objectives, eligibility criteria, sources of evidence, charting methods, results, and conclusions that relate to the review questions and objectives. | 2 |
| **INTRODUCTION** |  |  |  |
| Rationale | 3 | Describe the rationale for the review in the context of what is already known. Explain why the review questions/objectives lend themselves to a scoping review approach. | 3 |
| Objectives | 4 | Provide an explicit statement of the questions and objectives being addressed with reference to their key elements (e.g., population or participants, concepts, and context) or other relevant key elements used to conceptualize the review questions and/or objectives. | 4 |
| **METHODS** |  |  |  |
| Protocol and registration | 5 | Indicate whether a review protocol exists; state if and where it can be accessed (e.g., a Web address); and if available, provide registration information, including the registration number. | 4 |
| Eligibility criteria | 6 | Specify characteristics of the sources of evidence used as eligibility criteria (e.g., years considered, language, and publication status), and provide a rationale. | 4 |
| Information sources | 7 | Describe all information sources in the search (e.g., databases with dates of coverage and contact with authors to identify additional sources), as well as the date the most recent search was executed. | 4 |
| Search | 8 | Present the full electronic search strategy for at least 1 database, including any limits used, such that it could be repeated. | 28 |
| Selection of sources of evidence | 9 | State the process for selecting sources of evidence (i.e., screening and eligibility) included in the scoping review. | 5 |
| Data charting process | 10 | Describe the methods of charting data from the included sources of evidence (e.g., calibrated forms or forms that have been tested by the team before their use, and whether data charting was done independently or in duplicate) and any processes for obtaining and confirming data from investigators. | 6 |
| Data items | 11 | List and define all variables for which data were sought and any assumptions and simplifications made. | 7 |
| Critical appraisal of individual sources of evidence | 12 | If done, provide a rationale for conducting a critical appraisal of included sources of evidence; describe the methods used and how this information was used in any data synthesis (if appropriate). | 7 |
| Synthesis of results | 13 | Describe the methods of handling and summarizing the data that were charted. | 7 |
| **RESULTS** |  |  |  |
| Selection of sources of evidence | 14 | Give numbers of sources of evidence screened, assessed for eligibility, and included in the review, with reasons for exclusions at each stage, ideally using a flow diagram. | 8 |
| Characteristics of sources of evidence | 15 | For each source of evidence, present characteristics for which data were charted and provide the citations. | 9 – 10 |
| Critical appraisal within sources of evidence | 16 | If done, present data on critical appraisal of included sources of evidence (see item 12). | 19 |
| Results of individual sources of evidence | 17 | For each included source of evidence, present the relevant data that were charted that relate to the review questions and objectives. | 15 – 17 |
| Synthesis of results | 18 | Summarize and/or present the charting results as they relate to the review questions and objectives. | 11 – 14 |
| **DISCUSSION** |  |  |  |
| Summary of evidence | 19 | Summarize the main results (including an overview of concepts, themes, and types of evidence available), link to the review questions and objectives, and consider the relevance to key groups. | 19 |
| Limitations | 20 | Discuss the limitations of the scoping review process. | 21 |
| Conclusions | 21 | Provide a general interpretation of the results with respect to the review questions and objectives, as well as potential implications and/or next steps. | 21 |
| **FUNDING** |  |  |  |
| Funding | 22 | Describe sources of funding for the included sources of evidence, as well as sources of funding for the scoping review. Describe the role of the funders of the scoping review. | 21 |

**Table A2.**
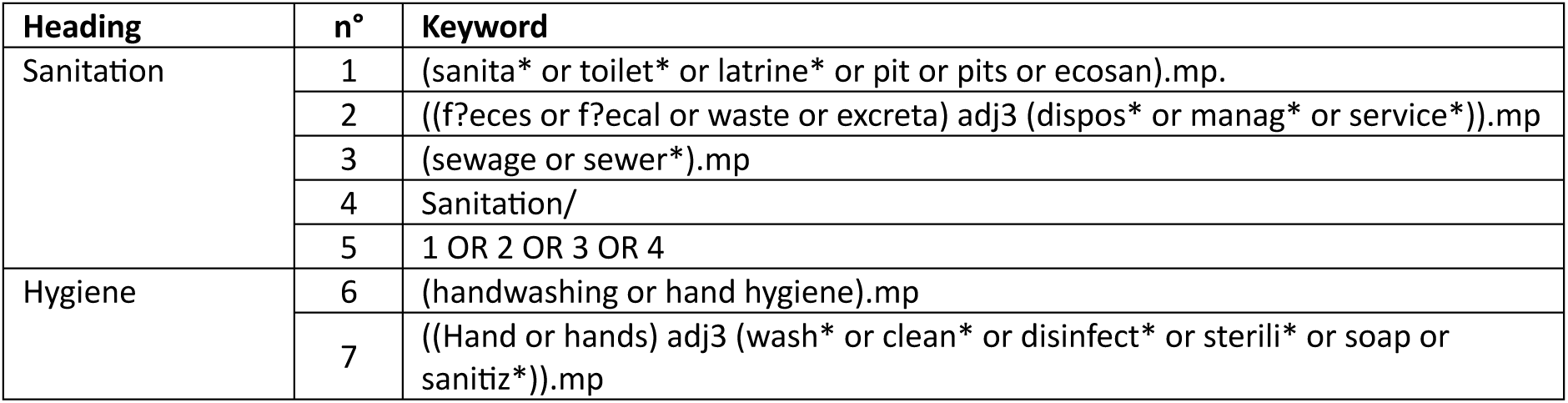
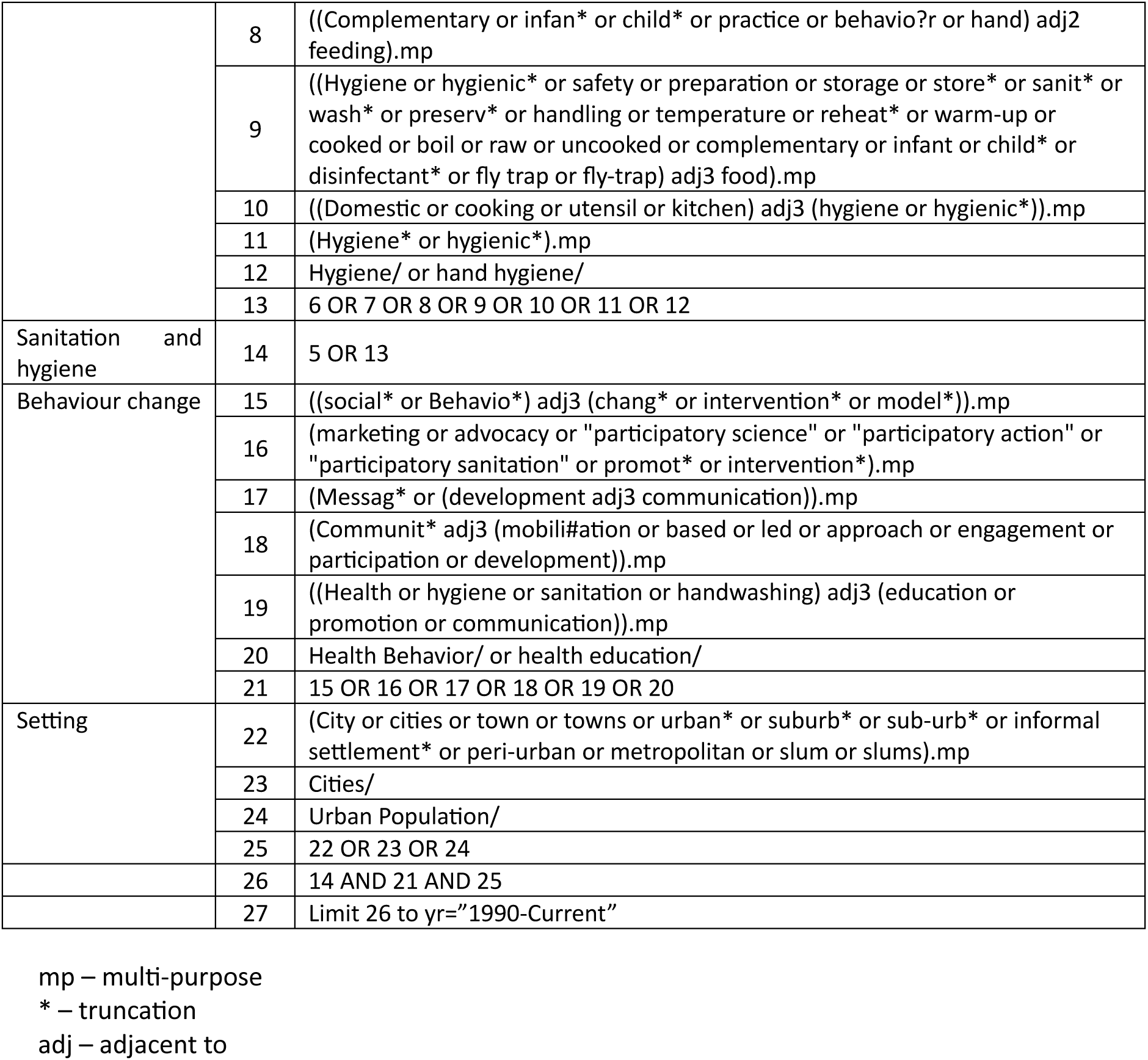
Search terms.

**Table A3.**
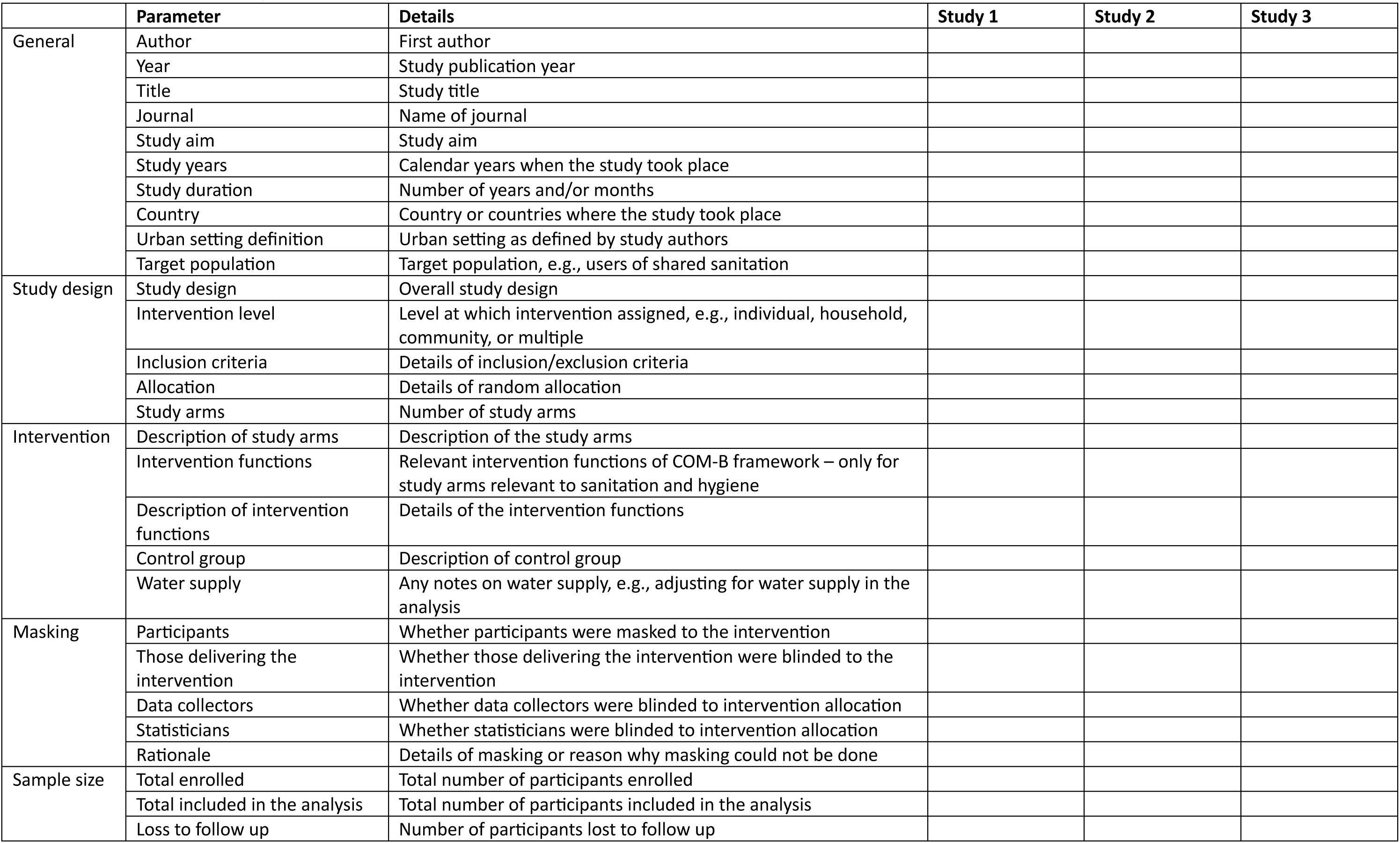
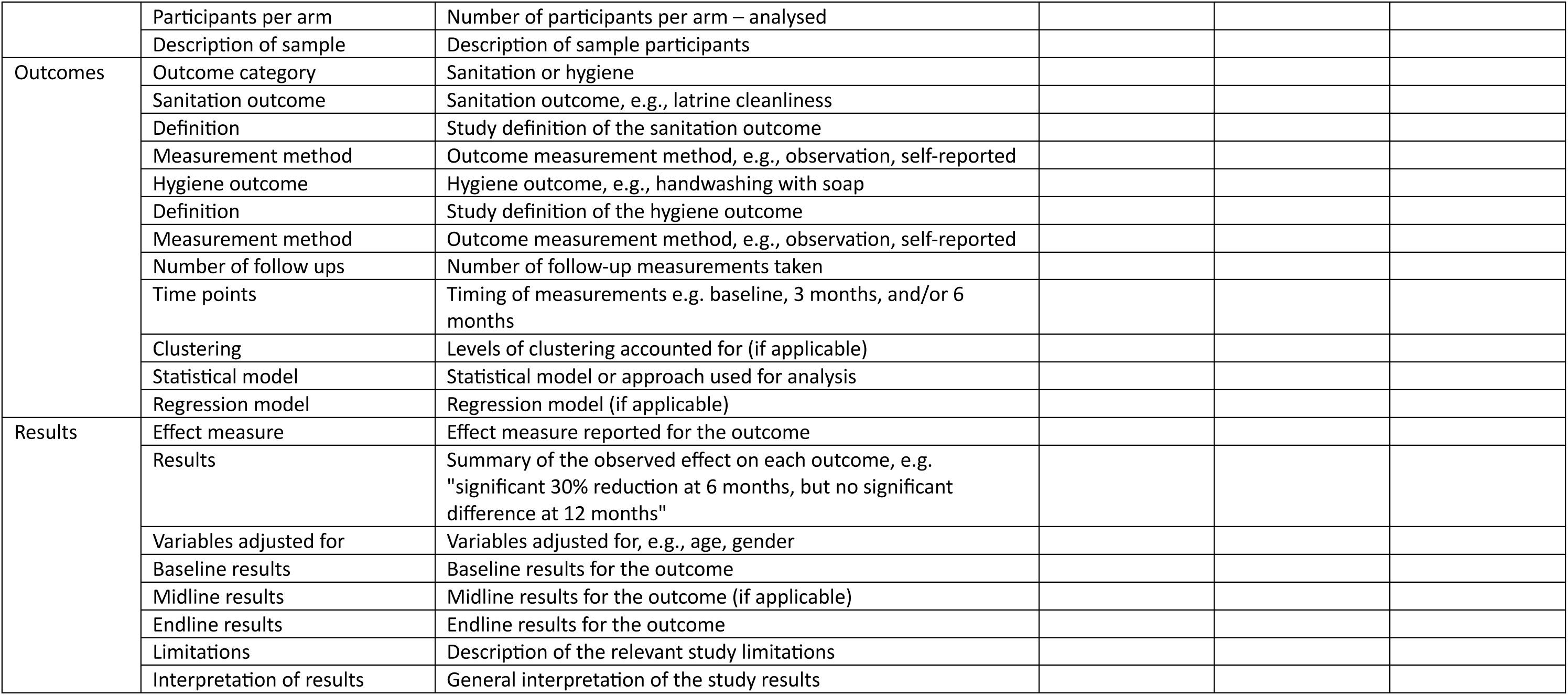
Data extraction template.

**Table A4.** Risk of bias scoring.

| Criterion | Question | Scoring guidance | Maximum score |
| --- | --- | --- | --- |
| Selection bias | Is there evidence of selection bias, which refers to systematic differences between baseline characteristics of the groups that are compared? | RCTs/cRCTs receive 1 star, unless evidence of selection bias (e.g., randomisation procedures not followed). Meaningful differences between groups at baseline in (c)RCTs receive 0 stars. Rates of declining to participate >10% (or not reported) receive 0 stars. Non- or quasi-randomised studies receive 0 stars. | 1 |
| Response bias | Is there evidence of response bias? | If intervention recipient was not blinded to intervention status, 0 stars. | 1 |
| Follow up bias | Is there evidence of bias due to missing follow-up data? | <10% receives 1 star, ≥10% receives 0 stars, not reported receives 0 stars. | 1 |
| Misclassification bias | Is there risk of households not receiving the intervention being misclassified as having received it, or vice versa? | Interventions delivered to specific households/individuals receive 1 star. Interventions delivered to communities in general that missed a substantial, i.e. ≥10%, proportion of the target population receive 0 stars, including when there is insufficient information to verify whether this is the case. Interventions with substantial risk of contamination (control households receiving intervention) receive 0 stars. | 1 |
| Outcome assessment | Is there evidence of bias arising from how the outcome was assessed? | Parent / person recall (=0 stars). Fieldworker assessed (=1 star). Physician / microbiologically assessed (=2 stars) | 2 |
| Outcome measurement | Is there evidence of ascertainment bias? | If outcome measurement staff were not blinded to intervention status, 0 stars. | 1 |
| Bias in analysis | Is there evidence that analysis was not appropriately adjusted for clustering and/or confounding, if appropriate? | Scoring is based on losing stars (max. 2). Individual RCTs with baseline balance on covariates are unlikely to require adjustment (=2 stars). Cluster-RCTs and non-randomised trials may require adjustment for clustering (-1 star if not done). RCTs or cRCTs may require adjustment for covariates, with justification (-1 star if adjustment appeared necessary but was not done). Non-randomised studies require adjustment for covariates (-1 star if not done), but also adequate justification for covariate selection (-1 star if not included). | 2 |

**Table A4.** List of excluded documents with reasons.

| Author | Year | Title | Exclusion reason |
| --- | --- | --- | --- |
| Aluko et al. | 2018 | The dynamics and determinants of household shared sanitation cleanliness in a heterogeneous urban settlement in Southwest Nigeria | 3 |
| Asamane et al. | 2023 | Protocol for a parallel group, two-arm, superiority cluster randomised trial to evaluate a community-level complementary-food safety and hygiene and nutrition intervention in Mali: the MaaCiwara study | 1 |
| Ashraf et al. | 2021 | Design and rationale of the Longitudinal Evaluation of Norms and Networks Study (LENNS): A cluster-randomized trial assessing the impact of a norms-centric intervention on exclusive toilet use and maintenance in peri urban communities of Tamil Nadu | 1 |
| Asrate et al. | 2022 | Households' access to an improved latrine and its associated factors among households of sanitation marketing products users and non-users, Northeast Amhara, Ethiopia | 3 |
| Bicchieri et al. | 2020 | The Norms and Networks Sanitation Study in India in Tamil Nadu, India | 1 |
| Biran et al. | 2011 | Patterns and determinants of communal latrine usage in urban poverty pockets in Bhopal, India. Trop Med Int Health 2011; 16:854–862 | 6 |
| Biran et al. | 2014 | Effect of a behaviour-change intervention on handwashing with soap in India (SuperAmma): a cluster-randomised trial | 5 |
| Cairncross et al. | 2005 | What causes sustainable changes in hygiene behaviour? A cross-sectional study from Kerala, India | 4 |
| Capone et al. | 2020 | Impact of an intervention to improve pit latrine emptying practices in low income urban neighborhoods of Maputo, Mozambique | 3 |
| Chase et al. | 2012 | Handwashing Behavior Change at Scale: Evidence from a Randomized Evaluation in Vietnam | 5 |
| Cole et al. | 2015 | Exploring the utility of diffusion theory to evaluate social marketing approaches to improve urban sanitation in Malawi | 3 |
| Contzen et al. | 2015 | Changing handwashing behaviour in southern Ethiopia: a longitudinal study on infrastructural and commitment interventions | 5 |
| Cookey et al. | 2020 | Perception management of non-sewered sanitation systems towards scheduled faecal sludge emptying behaviour change intervention | 3 |
| Curtis et al. | 2001 | Evidence of behaviour change following a hygiene promotion programme in Burkina Faso | 4 |
| Delea et al. | 2019 | Design of a parallel cluster-randomized trial assessing the impact of a demand-side sanitation and hygiene intervention on sustained behavior change and mental well-being in rural and peri-urban Amhara, Ethiopia: Andilaye study protocol | 3 |
| Feachem | 1984 | Interventions for the control of diarrhoeal diseases among young children: promotion of personal and domestic hygiene | 4 |
| Freeman et al. | 2022 | The impact of a demand-side sanitation and hygiene promotion intervention on sustained behavior change and health in Amhara, Ethiopia: A cluster-randomized trial | 5 |
| Galiani | 2012 | Promoting Handwashing Behavior in Peru: The Effect of Large-Scale Mass-Media and Community Level Interventions | 5 |
| Gautam et al. | 2017 | Trial of a novel intervention to improve multiple food hygiene behaviors in Nepal | 5 |
| Greenland et al. | 2016 | Multiple behaviour change intervention for diarrhoea control in Lusaka, Zambia: a cluster randomised trial | 4 |
| Grover et al. | 2018 | Comparing the behavioural impact of a nudge-based handwashing intervention to high-intensity hygiene education: a cluster-randomised trial in rural Bangladesh | 5 |
| Halcrow et al. | 2014 | Developing behaviour change communication for improving faecal sludge management in Bhutan | 3 |
| Hermida et al. | 2019 | Handwashing promotion in 10-year-old children: intervention studies in Northern Argentina | 8 |
| Hulland et al. | 2013 | Designing a handwashing station for infrastructure-restricted communities in Bangladesh using the integrated behavioural model for water, sanitation and hygiene interventions (IBM-WASH) | 3 |
| Jenkins & Scott | 2007 | Behavioural indicators of household decision-making and demand for sanitation and potential gains from social marketing in Ghana | 6 |
| Jenkins et al. | 2015 | Pit latrine emptying behavior and demand for sanitation services in dar Es Salaam, Tanzania | 3 |
| Jongpiputvanich, Veeravongs, & Wonsekiarttira | 1998 | Difficulties in conducting participatory action research to prevent diarrhoea in a slum area of Bangkok | 6 |
| Kaewchana et al. | 2012 | Effect of intensive hand washing education on hand washing behaviors in Thai households with an influenza-positive child in urban Thailand | 5 |
| Lagerkvist et al. | 2014 | Health in perspective: framing motivational factors for personal sanitation in urban slums in Nairobi, Kenya, using anchored best-worst scaling | 3 |
| Langford, Lunn, & Panter-Brick | 2011 | Hand-washing, subclinical infections, and growth: A longitudinal evaluation of an intervention in Nepali slums | 7 |
| Legge et al. | 2022 | Urban Water Access and Use in the Kivus: Evaluating Behavioural Outcomes Following an Integrated WASH Intervention in Goma and Bukavu, Democratic Republic of Congo | 7 |
| Luby et al. | 2014 | Using Behaviour Change Messaging to Improve Communal Toilets in Dhaka, Bangladesh | 2 |
| Luby et al. | 2006 | Combining drinking water treatment and hand washing for diarrhoea prevention, a cluster randomised controlled trial | 7 |
| Luby et al. | 2001 | Microbiologic effectiveness of hand washing with soap in an urban squatter settlement, Karachi, Pakistan | 7 |
| Luby et al. | 2004 | Effect of intensive handwashing promotion on childhood diarrhea in high-risk communities in Pakistan: A randomized controlled trial | 7 |
| Manjang et al. | 2018 | Promoting hygienic weaning food handling practices through a community- based programme: intervention implementation and baseline characteristics for a cluster randomised controlled trial in rural Gambia | 5 |
| Monte et al. | 1997 | Designing educational messages to improve weaning food hygiene practices of families living in poverty | 4 |
| Mosler & Sonego | 2017 | Improved latrine cleanliness through behaviour change and changes in quality of latrine construction: a longitudinal intervention study in rural Burundi | 5 |
| Musoke et al. | 2018 | Drinking water supply, sanitation, and hygiene promotion interventions in two slum communities in Central Uganda | 4 |
| Nakarutimana et al. | 2022 | Evaluation of the Control of Water Sanitation and Hygiene-Related Disease Through Community Hygiene Club Intervention in Rwanda | 3 |
| Ndaw | 2016 | Unlocking the Potential of Information Communications Technology to Improve Water and Sanitation Services | 3 |
| Obika et al. | 2006 | Creating demand for urban sanitation in sub-Saharan Africa | 3 |
| Peletz et al. | 2020 | Expanding safe fecal sludge management in Kisumu, Kenya: an experimental comparison of latrine pit-emptying services | 3 |
| Pinfold & Horan | 1996 | Measuring the effect of a hygiene behaviour intervention by indicators of behaviour and diarrhoeal diseases | 5 |
| Rahman et al. | 2017 | Behavioral antecedents for handwashing in a low-income urban setting in Bangladesh: an exploratory study | 3 |
| Ryman et al. | 2012 | Integration of routine vaccination and hygiene interventions: a comparison of 2 strategies in Kenya | 6 |
| Saxton et al. | 2017 | If I do not have enough water, then how could I bring additional water for toilet cleaning?! Addressing water scarcity to promote hygienic use of communal toilets in Dhaka, Bangladesh | 3 |
| Sequeira et al. | 2019 | Assessing the short-term outcomes of a piped water supply intervention in peri-urban Mozambique | 6 |
| Shapu et al. | 2021 | Effectiveness of Health Education Intervention on Water Sanitation and Hygiene Practice among Adolescent Girls in Maiduguri Metropolitan Council, Borno State, Nigeria: A Cluster Randomised Control Trial | 5 |
| Shiras et al. | 2018 | Shared sanitation management and the role of social capital: findings from an urban sanitation intervention in Maputo, Mozambique | 6 |
| Simiyu et al. | 2020a | Barriers and opportunities for cleanliness of shared sanitation facilities in low-income settlements in Kenya | 3 |
| Simiyu et al. | 2020b | Designing a food hygiene intervention in low-income, peri-urban context of Kisumu, Kenya: application of the trials of improved practices methodology | 3 |
| Simiyu et al. | 2021 | Developing and testing strategies for improving cleanliness of shared sanitation in low-income settlements of Kisumu, Kenya | 3 |
| Stanton & Clemens | 1987 | An educational intervention for altering water-sanitation behaviors to reduce childhood diarrhea in urban Bangladesh. II. A randomized trial to assess the impact of the intervention on hygienic behaviors and rates of diarrhea | 9 |
| Stanton et al. | 1992 | Theory-driven behavioral intervention research for the control of diarrheal diseases | 6 |
| Takanashi et al. | 2013 | Long-term impact of community-based information, education and communication activities on food hygiene and food safety behaviors in Vietnam: a longitudinal study | 4 |
| Tidwell et al. | 2018 | Sanitation Demand Creation in Peri-Urban Slums of Lusaka, Zambia | 3 |
| Tidwell et al. | 2019b | Theory-driven formative research on on-site, shared sanitation quality improvement among landlords and tenants in peri-urban Lusaka, Zambia | 3 |
| Tidwell et al. | 2019c | Understanding demand for higher quality sanitation in peri-urban Lusaka, Zambia through stated and revealed preference analysis | 3 |
| Tidwell et al. | 2019d | Using a theory-driven creative process to design a peri-urban on-site sanitation quality improvement intervention | 3 |
| Tumwebaze | 2014 | Shared toilet users' collective cleaning and determinant factors in Kampala slums, Uganda | 3 |
| USAID | 2011 | Pakistan safe drinking water and hygiene promotion project: evaluation report | 5 |
| Whaley & Webster | 2011 | The effectiveness and sustainability of two demand-driven sanitation and hygiene approaches in Zimbabwe | 5 |
| Whittington et al. | 1993 | Household demand for improved sanitation services in Kumasi, Ghana: a contingent valuation study | 9 |
| Yang et al. | 2017 | Effectiveness of a multifaceted intervention on improving the hand-washing skills and behaviors of migrant workers in Beijing | 5 |
| Zisa et al. | 2022 | Achieving handwashing with social art for a behavioural change: the experience of the Lazos de Agua programme in Latin America | 4 |

1= protocol; 2= clinical trial registration; 3= study design; 4= no control group; 5= not in an urban setting; 6= no behaviour change intervention; 7= no sanitation or hygiene outcome; 8= not published in English; 9= published before 1 January 1990

**Table A5.** Risk of bias scoring of included studies.

| Study | Selection bias | Response bias | Follow-up bias | Misclassification bias | Outcome assessment | Outcome measurement | Bias in analysis | Total score (max 9) |
| --- | --- | --- | --- | --- | --- | --- | --- | --- |

| Sanitation |  |  |  |  |  |  |  |  |
| --- | --- | --- | --- | --- | --- | --- | --- | --- |
| Alam et al. (16) | 1- cRCT | 0- not blinded | 1- <10% loss to follow up | 1- household level | 1- fieldworker observation | 1- yes | 2- cluster adjusted difference | 7 |
| Bick et al. (14) | 0- not randomised | 0- not blinded | 1- <10% loss to follow up | 1- compound level | 1- fieldworker observation | 0- not stated | 1- non-independence within compounds | 4 |
| Tidwell et al. (24) | 1- RCT | 0- not blinded | 0- >10% loss to follow up (14%) | 1- individual level | 1- fieldworker observation | 1- yes | 2- monthly rent, landlord education level, presence of a separate toilet for the landlord, presence of water on the plot, and the number of households living on the plot | 4 |
| Tumwebaze et al. (25) | 0- not randomised | 0- not blinded | 0- >10% loss to follow up (70%) | 1- individual level | 0- structured household questionnaire | 1- yes | 0- not stated | 2 |
| Yeager et al. (26) | 0- not randomised | 0- not blinded | 1- <10% loss to follow up | 1- individual level | 1- fieldworker observation | 0- not stated | 0- not stated | 3 |
| Hygiene |  |  |  |  |  |  |  |  |
| Amon-Tanoh et al. (17) | 1- cRCT | 0- not blinded | 1- <10% loss to follow up | 1- compound level | 1- fieldworker observation | 1- yes | 2- age, gender, grp, education, level of female head of household, and baseline handwashing with soap | 7 |
| Bick et al. (14) | 0- not randomised | 0- not blinded | 1- <10% loss to follow up | 1- compound level | 1- fieldworker observation | 0- not stated | 1- non-independence within compounds | 4 |
| Biswas et al. (18) | 1- cRCT | 0- not blinded | 1- <10% loss to follow up | 1- household level | 1- fieldworker observation | 0- not stated | 2- no mention of clustering | 6 |
| Davis et al. (20) | 0- not randomised | 0- not blinded | 0- >10% loss to follow up (25%) | 1- household level | 1- fieldworker observation | 0- not stated | 1- baseline differences | 2 |
| Guiteras et al. (21) | 1- RCT | 0- not blinded | 0- not reported | 1- compound level | 1- fieldworker observation | 0- not stated | 0- not stated | 3 |
| Simiyu et al. (15) | 1- process evaluation of RCT | 0- not blinded | 0- >10% loss to follow up (11%) | 1- individual level | 1- fieldworker observation | 1- yes | 0- not stated | 4 |
| Langford et al. (22) | 1- RCT | 0- not blinded | 1- <10% loss to follow up | 1- individual level | 1- lead author observation | 0- not stated | 0- not stated | 4 |
| Luby et al. (23) | 1- cRCT | 0- not blinded | 0- >10% loss to follow up (30%) | 1- household level | 1- fieldworker observation | 0- not stated | 2- clustering | 5 |
| Bowen et al. (19) | 1- cRCT | 0- not blinded | 0- >10% loss to follow up (18%) | 1- household level | 1- fieldworker observation | 0- no | 2- clustering | 5 |

## Notes

### Competing Interest Statement

The authors have declared no competing interest.

### Funding Statement

This research was funded by the United Kingdom Foreign, Commonwealth and Development Office (grant code: 301186). The funder had no role in study design, data collection and analysis, decision to publish, nor preparation of the manuscript.

